# Rate of Force Development is Related to Maximal Force, Muscle Quality and Physical Function in Patients With Chronic Kidney Disease Predialysis

**DOI:** 10.1101/19013557

**Authors:** Jared M. Gollie, Michael O. Harris-Love, Samir S. Patel, Marc R. Blackman

## Abstract

**Background:** Physical function is severely compromised in people with chronic kidney disease (CKD) and worsens with continued decreases in kidney function. Neuromuscular force capacity is a key determinant of physical function in healthy older adults, though its importance in persons with CKD is less understood.

**Methods:** This study aimed to determine the relationships among rate of force development (RFD), muscle quality and physical function in a group of community-dwelling, middle-aged and older men (n=14; age=71.2±6.2 years) with CKD stages 3 and 4 (eGFR=37.5±10.4 ml/min per 1.73 m^2^). Force characteristics were determined from maximal knee extensor isometric contractions and muscle quality was estimated using ultrasound grayscale analysis. Physical function was assessed by the Short Physical Performance Battery (SPPB) and 5-repetition sit-to-stand (STS) test.

**Results:** eGFR was directly related to SPPB (r=0.54, p=0.044) and inversely related to STS (r=-0.62, p=0.029). RFD was positively related to SPPB at time points 0-50 ms, 50-100 ms, and 0-300 ms (RFD_0-50_, r=0.73, p=0.010; RFD_50-100_, r=0.67, p=0.022 and RFD_0-300_ r=0.61, p=0.045); and inversely related to STS at time points 0-50 ms, 50-100 ms, and 0-300 ms (RFD_0-50_, r=-0.78, p=0.007; RFD_50-100_, r=-0.78, p=0.006 and RFD_0-300_ r=-0.76, p=0.009), respectively. RFD was positively associated with maximal voluntary force (MVF) at times 50-100 ms, 100-200 ms, and 0-300 ms (RFD_50-100_, r=0.72, p=0.011; RFD_100-200_, r=0.66, p=0.025; and RFD_0-300_ r=0.70, p=0.016), respectively. Neither MVF nor muscle quality was significantly associated with functional measures.

**Conclusions:** RFD is an important determinant of physical function in middle-aged and older men with CKD stages 3 and 4.

## INTRODUCTION

Physical function is severely compromised in people with chronic kidney disease (CKD) and has been shown to be predictive of risk of hospitalization, disability and mortality (10, 16, 17, 25, 26, 28, 30, 33). For example, individuals with low functional status at the time of dialysis initiation experience a higher risk of all-cause mortality irrespective of age and history of cardiovascular disease (10). Similarly, functional measures of gait speed and timed-up-and-go are better predictors of 3-year mortality than are kidney function or commonly measured serum biomarkers in patients with CKD predialysis (29).

Rate of force development (RFD) describes the rate at which force is expressed during fast and forceful contractions and has become recognized as an underlying factor contributing to physical function in older adults (8, 24). Early time intervals (i.e., <75 ms) of RFD are thought to be primarily influenced by neural factors, whereas muscular factors become increasingly important with greater time from onset of contraction (3, 20, 27). However, Gerstner et al. reported that age-associated declines in rapid torque production during later time intervals (i.e., ≥100 ms) are influenced by alterations in muscle quality, architecture, and muscle activation (4).

Accelerated skeletal muscle loss is a major concern in patients with CKD and end-stage renal disease (ESRD) (34). Reductions in muscle cross-sectional area have been associated with declines in physical function (13, 21). Neurological complications, including peripheral neuropathy, are also highly prevalent in patients with CKD (1, 15). Accordingly, both neural and muscle morphological alterations are potential factors contributing to muscle weakness. The loss in neuromuscular force is reported to occur more rapidly than changes in muscle cross-sectional area in patients with CKD predialysis and ESRD (14, 18). Moreover, changes in muscle cross-sectional area have been found to be poorly associated with changes in functional outcomes (14). Taken together, these observations further highlight the potential contributions of neural alterations, in addition to muscle morphological properties, on neuromuscular capacity in patients with CKD.

Currently, there is little information describing neuromuscular contributions to physical function in patients with CKD predialysis. Consequently, the effects of compromised force characteristics and muscle quality, if present, on physical function in those with CKD predialysis are unclear. In this study, we aimed to determine the relationships among RFD, muscle quality and physical function in patients with CKD stages 3 and 4 predialysis. We hypothesized that RFD, maximal voluntary force (MVF), and muscle quality would be significantly associated with physical function in patients with CKD stages 3 and 4.

## METHODS

### Study Design

A single cohort, cross-sectional study design was used to examine the relationships among knee extensor RFD, maximum voluntary force (MVF), muscle quality of the rectus femoris (RF), and sit-to-stand (STS) time in a convenience sample of middle-aged and older men with CKD stages 3 and 4 seen at a Veterans Affairs Medical Center. Data were collected by trained staff in the Renal Clinic, and Physical Medicine and Rehabilitation and Research Service groups at the Veterans Affairs Medical Center in Washington, DC (DC VAMC) and the study was registered with ClinicalTrials.gov (NCT03160326).

### Participants and Ethical Approval

Community-dwelling male Veterans were screened and referred by the Renal Clinic staff at the DC VAMC for potential enrollment. The study was approved by the DC VAMC Institutional Review Board and Research and Development Committee. Inclusion criteria for study enrollment required participants to be ambulatory, with or without use of an assistive device, aged 18-85 years, and diagnosed with CKD stage 3 or 4, based on eGFR measures obtained within the 3-months prior to study enrollment. Exclusion criteria included a history of acute kidney injury, any uncontrolled cardiovascular or musculoskeletal problems, or having a pacemaker or implantable cardioverter defibrillator. All participants voluntarily provided written informed consent using an IRB approved form prior to study participation.

### Procedures

Each participant reported to the Skeletal Muscle Laboratory in the Clinical Research Center at the DC VAMC to complete neuromuscular strength, muscle quality and physical function assessments. Participants were asked to refrain from any vigorous activity for at least 24 hours before testing. Quantitative ultrasound imaging preceded strength and functional testing. At least 30-minutes was allotted between strength and functional assessments to minimize the potential for test contamination due to fatigue.

### Diagnostic Ultrasound Assessments

Methods used for diagnostic ultrasound assessments have previously been published (9, 11). Sonographic images were obtained using B-mode diagnostic ultrasound with a 5-18 MHz linear array transducer (Nobulus, Hitachi Aloka Medical, Parsippany, NJ). Manufacturer default settings for time gain compensation and near field/far field gain were applied for all musculoskeletal scans. The field of view was adjusted for each participant to optimally capture the region of interest (ROI) of the RF on the dominant side. Standardized procedures were followed for the identification of the scanning site, as previously described (9, 11). Scanned images were obtained by a single clinical investigator (MHL) with more than 10 years of quantitative ultrasound experience.

### Quantitative Strength Testing

Knee extensor strength of the dominant leg was assessed using a portable fixed dynamometer (FGV-200XY, NIDEC instruments, Itasca, IL). The dynamometer was securely mounted and anchored behind the participant. Participants were placed in the seated position on the edge of an adjustable examination table and with their hips and knees at 90° of flexion. A strap attached to the dynamometer was secured around the ankle of the dominant leg. Participants were allowed to use their hands for stability but were asked to refrain from grasping onto the table. A familiarization session was provided before data collection to orient each participant to the isometric testing procedures. Participants were instructed to extend or kick their leg as “hard” as possible following the test administrator’s requests, and to contract and sustain the contraction for 5-seconds while being provided strong verbal encouragement. A rest period of approximately 1-minute was provided between repetitions. Pre-contraction tension and countermovement were removed by test administrators and the dynamometer was zeroed just before contraction initiation.

### Short Physical Performance Battery (SPPB)

The SPPB is a measure used to assess lower extremity function and is predictive of subsequent disability in persons over 70 years of age (6). The SPPB is composed of tasks assessing balance, walking speed, and the ability to rise from a chair. A composite score is generated with the highest possible score being 12. Those completing the respective tasks were assigned a score of 1 to 4 on each task, with higher scores representative of better performance of the task. Participants unable to complete a task were given a score of zero for that task. Complete details of the SPPB have been described elsewhere (7).

### Sit-to-Stand (STS)

For the purposes of this study we also examined the STS task independent of the SPPB as the ability to rise from a chair repeatedly has been used as a measure to assess lower extremity power (32). STS determined as the time taken to complete 5 sit-to-stand repetitions. The test was stopped if the participant became tired or short of breath, used his arms, was unable to complete the five repetitions within one-minute, or if the test administrator became concerned about the participant’s safety.

### Data analysis

#### Quantitative Ultrasound

All scanned images were obtained and measured 3 times within the fascial borders of the muscle. The ROI selection and echogenicity measures of the RF via grayscale (GSL) histogram analysis were completed using ImageJ (version 1.48; National Institutes of Health, Bethesda, MD, USA). ROI was defined as the area within the superior and inferior fascial borders and the lateral borders of the RF. In those instances when a portion of a fascial border was poorly visualized, the examiner used the trajectory of the visible fascial border to complete the ROI selection (11). Subcutaneous fat was quantified according to methods described by Stock et al. (31). The mean of the three thickness values from each image was utilized. GSL values were then corrected for subcutaneous fat using the following equation (31): corrected GSL = uncorrected GSL + (subcutaneous fat thickness [cm] x 40.5278).

#### Strength Testing

The real-time force applied to the dynamometer was displayed online on a computer monitor. Force data were sampled at 100Hz and digitally stored to be analyzed offline. No more than six maximal voluntary isometric contractions were completed during strength assessments. Peak isometric knee extensor strength was determined by averaging 3 attempts which fell within 10% of each other or the 3 highest values of the six attempts. Rate of force development (RFD) was calculated as the change in force by change in time following the initiation of isometric knee extension contraction and assessed over consecutive time epochs; 0-50 ms, 50-100 ms, 100-200 ms, and 200-300 ms (i.e., RFD_0-50_, RFD_50-100_, RFD_100-200_, and RFD_200-300_). RFD was also calculated as the change in force by the change in time over the first 300 ms of the contraction (i.e., RFD_0-300_) (**Figure 1**). The onset of contraction was determined as the first derivative of the filtered force signaling that crossed 2.5% of peak force (20). In addition to absolute RFD, RFD is expressed relative to MVF (RFD/MVF), corrected RF GSL (RFD/GSL), and RF muscle thickness (MT) (RFD/MT) (19, 20, 27). Specific force was calculated as MVF divided by RF MT and absolute MVF was normalized to body mass (BM) to power two-thirds (N/kg^0.67^) (12).

**Figure 1.**
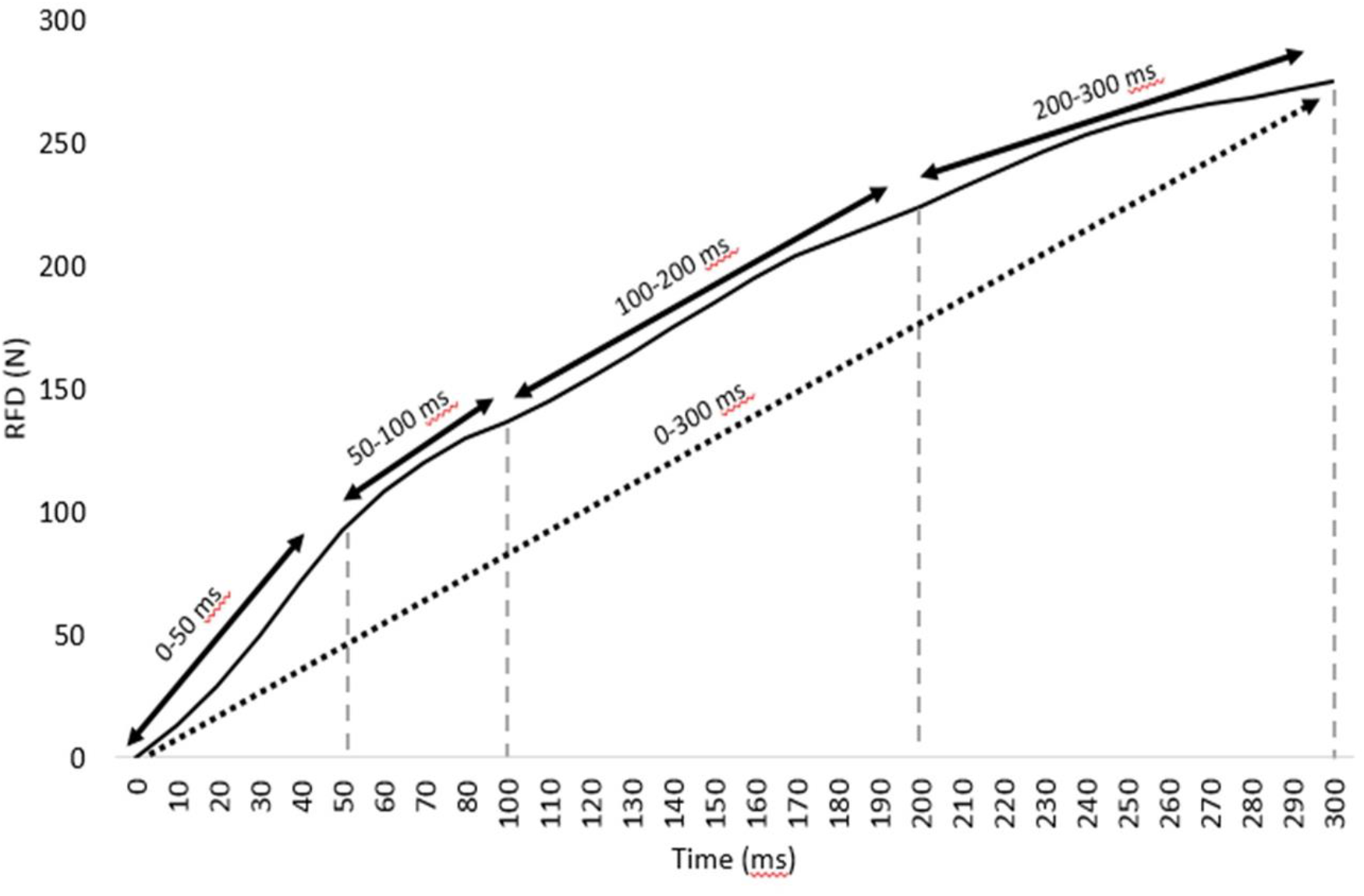
Example of a force-time curve during maximal voluntary isometric contraction of the dominant knee extensor depicting the specific time intervals used for assessment. *N*, newton; *ms*, millisecond

### Statistical analyses

Data are presented as mean values ± SD unless otherwise noted. Normality of each variable was assessed with the Shapiro-Wilk test. Pearson product-moment correlation coefficients (r) were used to determine associations among estimates of knee extensor strength, muscle quality, and physical function. The magnitudes of the correlation coefficients were described as 0.00-0.30, 0.30-0.50, 0.50-0.70, 0.70-0.90, and 0.90-1.00 and interpreted as negligible, low, moderate, high, and very high, respectively (23). Statistical significance was set at p<0.05 for two-tailed tests. Statistical analyses were performed using SPSS version 26 (SPSS Inc., Chicago, IL).

## RESULTS

Participant characteristics are presented in **Table 1**. Study participants were classified as exhibiting moderately to severely decreased kidney function based on mean eGFR, with 14% of participants classified with CKD stage 3a (n=2), 57% with CKD stage 3b (n=8), and 29% with CKD stage 4 (n=4). According to BMI, 36% of participants were classified as overweight (n=5) while 64% were classified as obese (n=9). Two of the fourteen participants, both classified as CKD stage 4, were unable to complete a single STS repetition and therefore were not included in the STS analysis. eGFR was directly related to the SPPB (r=0.54, p=0.044) and inversely related to STS (r=-0.62, p=0.029). No other significant relationships were observed between eGFR and RFD outcomes, MVF, RF GSL, or RF MT (data not shown). RFD data were unavailable on three participants; and therefore, eleven participants were included in the analysis of the relationships between physical function and muscle-specific outcomes with RFD unless otherwise noted. Of the three participants without RFD data, one was not able to complete the STS task.

**Table 1.** Participant characteristics (n=14).

| **Outcome** | **mean (SD)** |
| --- | --- |
| **Age (years)** | 71.2 (6.2) |
| **Height (cm)** | 175.1 (7.3) |
| **Weight (kg)** | 103.1 (22.1) |
| **BMI (kg/m<sup>2</sup>)** | 33.5 (5.9) |
| **eGFR (mL/min per 1.73 m<sup>2</sup>)** | 37.5 (10.4) |
| **Potassium (mmol/L)** | 4.7 (0.6) |
| **SPPB** | 8.6 (2.4) |
| **STS (s)\*** | 14.8 (3.3) |
| **MVF (N)** | 371 (78.8) |
| **Normalized MVF (N/kg<sup>0.67</sup>)** | 16.9 (3.7) |
| **Specific force (N/cm)** | 16.2 (5.1) |
| **RF GSL (a.u.)** | 80.4 (10.8) |
| **Corrected RF GSL (a.u.)** | 132.7 (19.7) |
| **RF MT (cm)** | 23.9 (5.0) |
| **Subcutaneous fat tissue (cm)** | 1.3 (0.5) |

\*n=12 for sit-to-stand (STS) task.
Abbreviations. *eGFR*, estimated glomerular filtration rate; *cm*, centimeters; *kg*, kilogram; *m*, meter; *mg*, milligram; *dl*, deciliter; *g*, gram; *%*, percent; *SPPB*, Short Physical Performance Battery; *STS*, sit-to-stand; *s*, seconds; *MVF*, maximal voluntary force; *N*, Newton; *RF GSL*, rectus femoris grayscale; *a.u.*, arbitrary units; *RF MT*, rectus femoris muscle thickness

### Short Physical Performance Battery (SPPB)

Participant RFD estimations are displayed in **Table 2. Figure 2** shows the relationships between the SPPB and RFD. SPPB was positively associated with RFD at time points 0-50 ms, 50-100 ms, and 0-300 ms (RFD_0-50_, r=0.73, p=0.010; RFD_50-100_, r=0.67, p=0.022 and RFD_0-300_r=0.61, p=0.045); and with RFD/GSL at time points 0-50 ms, 50-100 ms, and 0-300 ms (RFD/GSL_0-50_, r=0.70, p=0.015; RFD/GSL_50-100_, r=0.76, p=0.006 and RFD/GSL_0-300_ r=0.63, p=0.035), respectively. The SPPB was not significantly related to MVF, RF GLS, corrected RF GSL, RF MT, or specific force (data not shown).

**Table 2.** Absolute and relative rate of force development (RFD) estimations in male Veterans with CKD stages 3 and 4 (n=11).

| **Outcome** | **0-50ms** | **50-100 ms** | **100-200 ms** | **200-300 ms** | **0-300 ms** |
| --- | --- | --- | --- | --- | --- |
| **RFD (N·s)** | 1334.7<br>(713.9) | 988.2<br>(334.9) | 727.9<br>(371.9) | 382.5<br>(226.5) | 874.8<br>(272.2) |
| **RFD/MVF (N·s·N)** | 3.5 (1.8) | 2.5 (0.6) | 1.8 (0.8) | 1.0 (0.7) | 2.2 (0.5) |
| **RFD/GSL (N·s·a.u.)** | 10.7 (6.8) | 7.7 (2.9) | 5.8 (3.4) | 2.9 (1.8) | 6.9 (2.8) |
| **RFD/MT (N·s·cm)** | 58.0 (35.9) | 43.4 (17.3) | 33.5 (21.8) | 17.3 (12.6) | 39.1 (16.5) |

Data presented as mean values (SD).
Abbreviations. *RFD*, rate of force development; *RFD/MVF*, rate of force development normalized to maximal voluntary force; *RFD/GSL*, rate of force development normalized to corrected rectus femoris grayscale; *RFD/MT*, rate of force development normalized to rectus femoris muscle thickness; *N*, Newton; *s*, second; *a.u.*, arbitrary unit; *cm*, centimeter.

**Figure 2.**
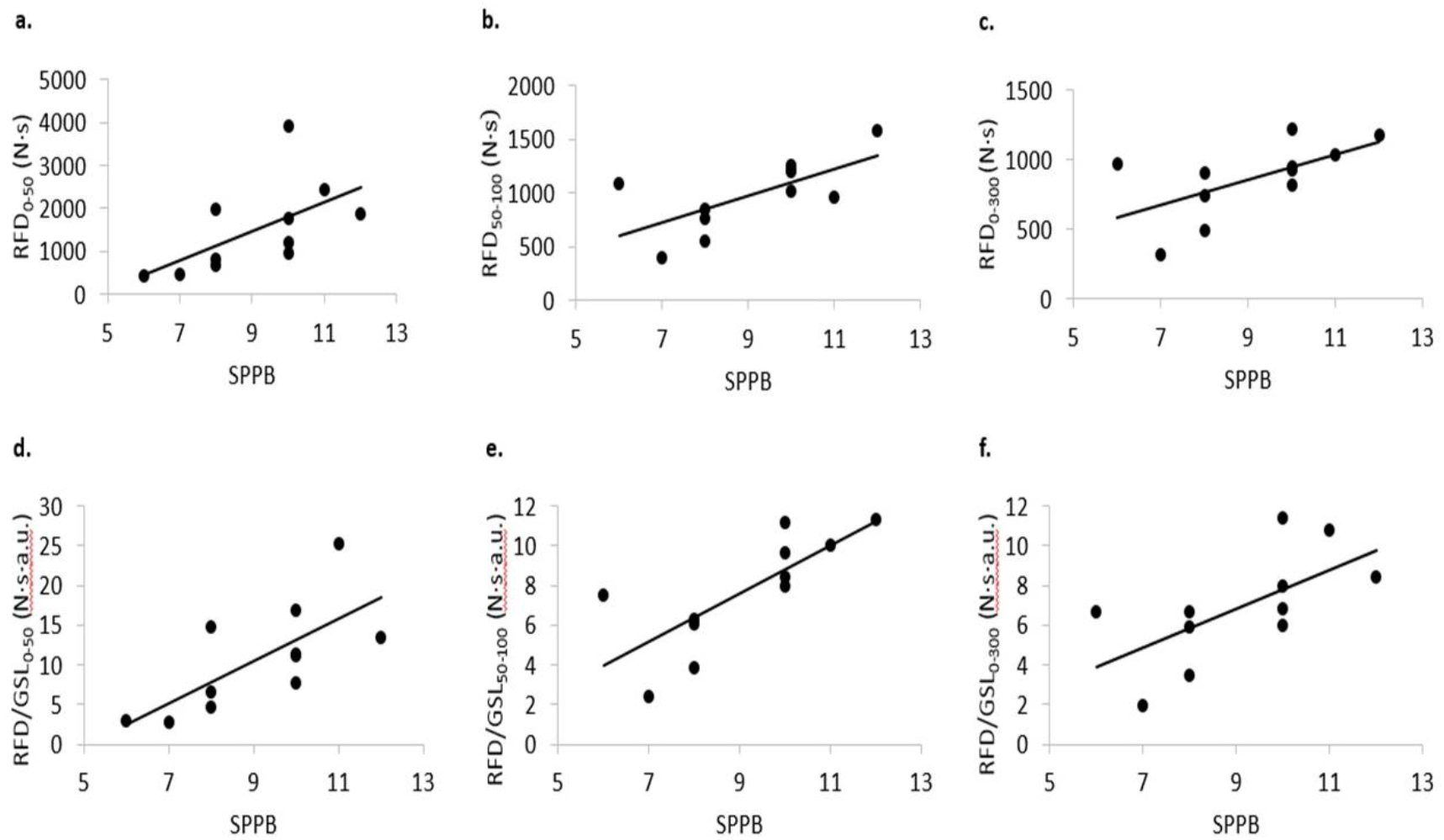
Relationship between SPPB and RFD at time intervals 0-50ms (a), 50-100 ms (b), and 0-300 ms (c); and the relationship between SPPB and RFD normalized to corrected RF GSL at time intervals 0-50 ms (d), 50-100 ms (e), and 0-300 ms (f). *RFD*, rate of force development; *GSL*, corrected rectus femoris grayscale; *SPPB*, Short Physical Performance Battery; *N*, Newton; *s*, second; *a.u*., arbitrary unit.

### Sit-to-Stand (STS)

Ten participants are included in the analysis of the relationship between STS and RFD due to two participants being unable to perform the STS assessment and the absence of RFD on three participants (**Figure 3**). The STS was inversely associated with RFD at time points 0-50 ms, 50-100 ms, and 0-300 ms (RFD_0-50_, r=-0.78, p=0.007; RFD_50-100_, r=-0.78, p=0.006 and RFD_0-300_ r=-0.76, p=0.009) and with RFD/MVF at time points 0-50 ms, 50-100 ms, and 0-300 ms (RFD/MVF_0-50_, r=-0.67, p=0.033; RFD/MVF_50-100_, r=-0.80, p=0.005 and RFD/MVF_0-300_ r=-0.73, p=0.015), respectively. The STS was also inversely related to RFD/GSL at time points 0-50 ms, 50-100 ms, and 0-300 ms (RFD/GSL_0-50_, r=-0.71, p=0.021; RFD/GSL_50-100_, r=-0.78, p=0.008 and RFD/GSL_0-300_ r=-0.64, p=0.042; respectively) and RFD/MT at time point 0-50 ms (RFD/MT_0-50_, r=-0.66, p=0.034), respectively. The STS was not significantly related to MVF, RF GSL, corrected RF GSL, RF MT, or specific force (data not shown).

**Figure 3.**
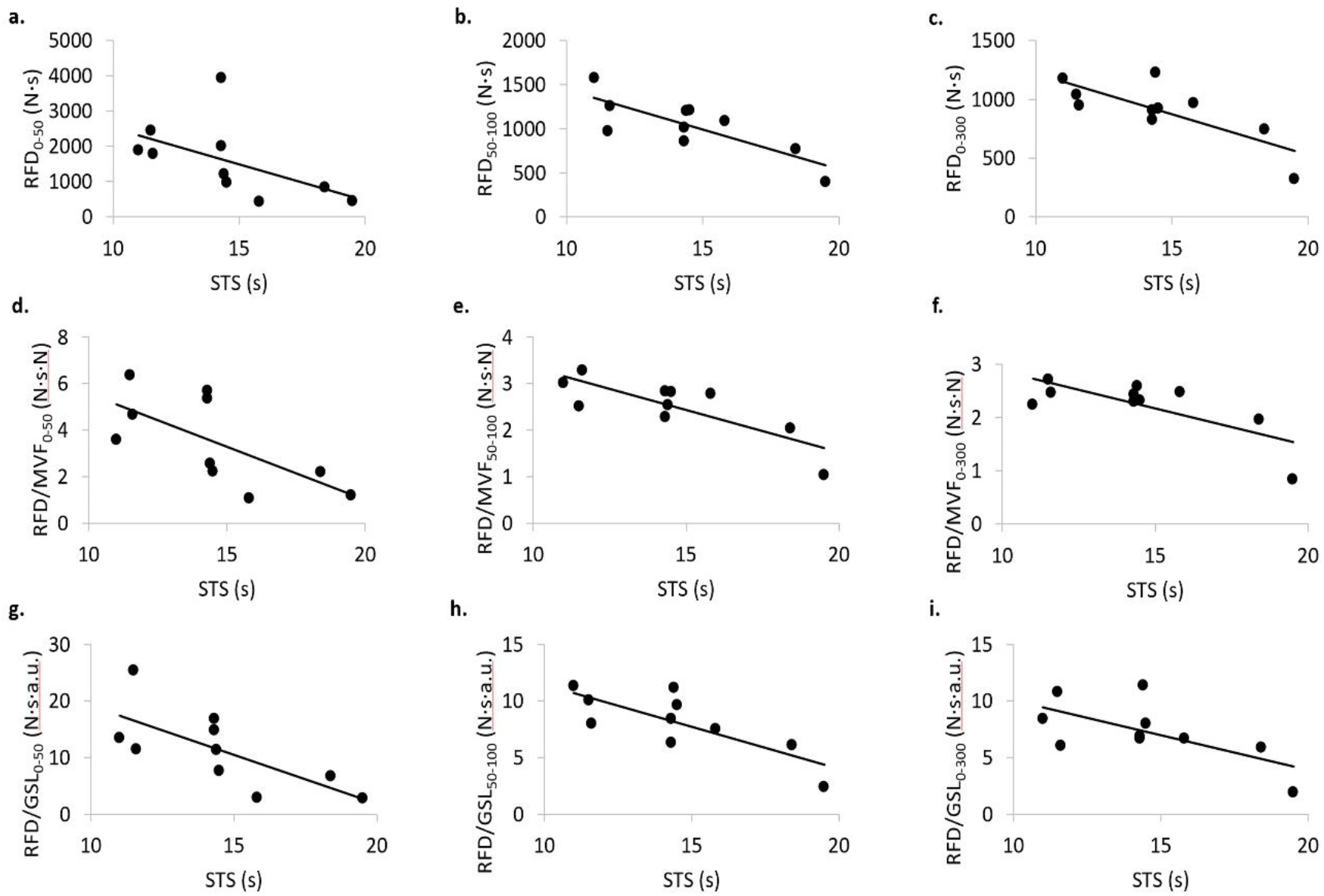
Relationship between STS and RFD at time intervals 0-50 ms (a), 50-100 ms (b), and 0-300 ms (c); the relationship between STS and RFD/MVF at time intervals 0-50 ms (d), 50-100 ms (e), and 0-300 ms (f); and the relationship between STS and RFD/GSL at time intervals 0-50 ms (g), 50-100 ms (h), and 0-300 ms (i). *RFD*, rate of force development; *MVF*, maximal voluntary force; *GSL*, corrected rectus femoris grayscale; *STS*, Sit-to-Stand; *N*, Newton; *s*, second; *a.u*., arbitrary unit.

### Maximal Voluntary Force (MVF)

**Figure 4** shows the relationships between the MVF and RFD. MVF was positively related to RFD at time points 50-100 ms, 100-200 ms, and 0-300 ms (RFD_50-100_, r=0.72, p=0.011; RFD_100-200_, r=0.66, p=0.025; and RFD_0-300_ r=0.70, p=0.016); and to RFD/GSL at time points 50-100 ms and 100-200 ms (RFD/GSL_50-100_, r=0.71, p=0.014 and RFD/GSL_100-200_, r=0.62, p=0.039) respectively; but not to RFD/MT (data not shown). Normalized MVF was positively related to RFD/GSL at time points 50-100 ms, 100-200 ms, and 0-300 (RFD/GSL_50-100_, r=0.61, p=0.042; RFD/GSL_100-200_, r=0.62, p=0.039; and to RFD/GSL_0-300_, r=0.64, p=0.034; respectively).

**Figure 4.**
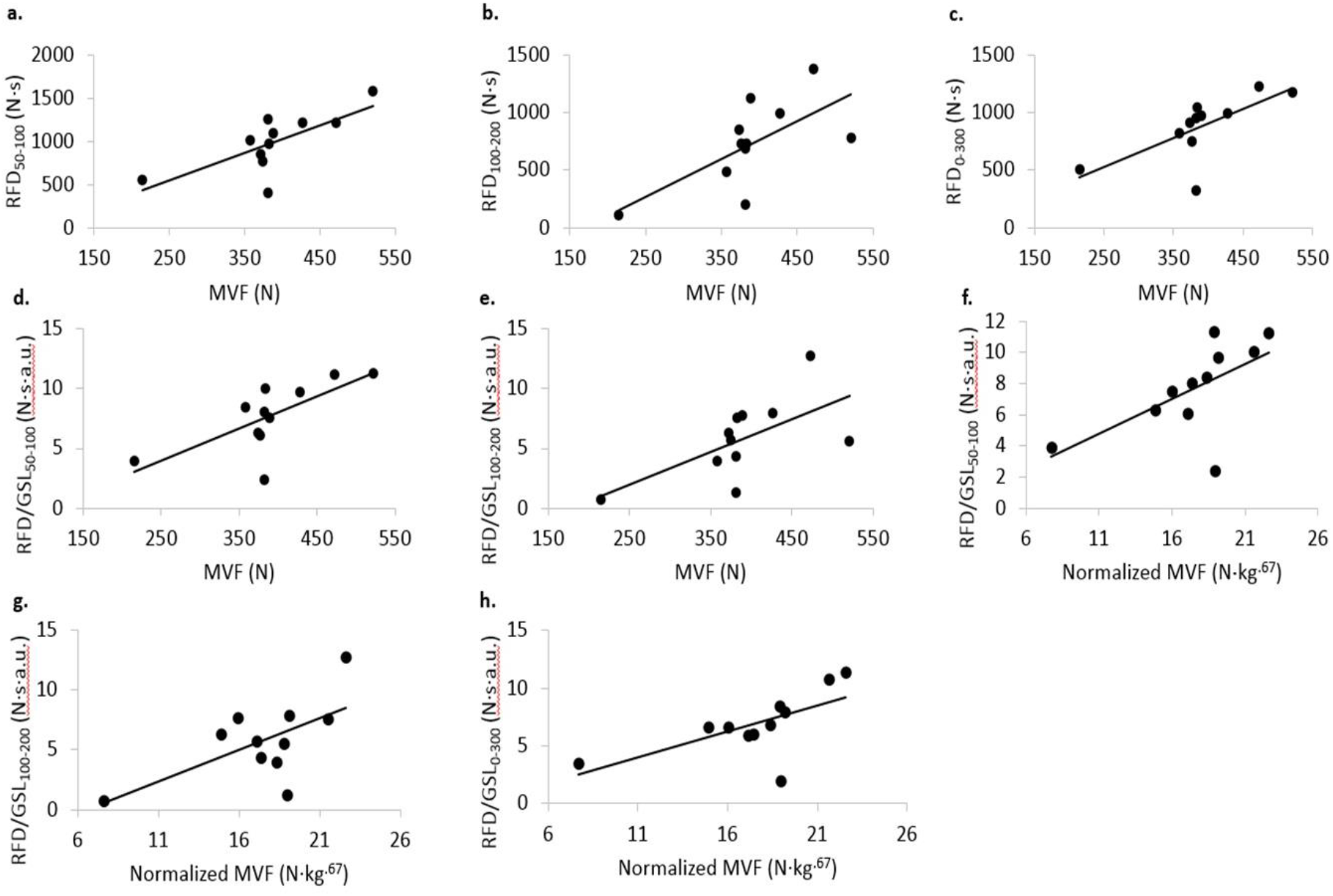
Relationship between RFD and MVF at time intervals 50-100 ms (a), 100-200 ms (b), and 0-300 ms (c); the relationship between RFD/GSL and MVF at time intervals 50-100 ms (d) and 100-200 ms; and the relationship between RFD/GSL and Normalized MVF at time intervals 50-100 ms (f), 100-200 ms (g), and 0-300 ms (h). *RFD*, rate of force development; *GSL*, corrected rectus femoris grayscale; *MVF*, maximal voluntary force. *N*, Newton; *kg*, kilogram; *s*, second; *a.u*., arbitrary unit.

### Muscle Quality

Corrected RF GSL was inversely associated with RFD/MVF at time point 0-300 (RFD/MVF_0-300_, r=-0.60; p=0.049) (*data not shown*).

### Specific Force

Specific force was positively associated with RFD at time point 100-200 ms (RFD_100-200_, r=0.67, p=0.022); and with RFD/GSL at time point 100-200 ms (RFD/GSL_100-200_, r=0.60, p=0.050) (*data not shown*).

## DISCUSSION

Reduced kidney function, as assessed by eGFR, was associated with declines in physical functional assessments. Moderate to strong relationships were found between RFD and SPPB, STS, and MVF. When controlling for MVF, RFD was moderately to strongly associated with STS and moderately related to muscle quality. While RFD was associated with MVF, no significant relationships were observed between measures of MVF and functional outcomes. The findings of the present study suggest that functional performance is significantly related to RFD in middle aged and older men with CKD stages 3 and 4 predialysis.

The decline in physical function is accelerated in patients with CKD predialysis and ESRD and progressively worsens as kidney function declines (16, 21). Variations of the STS task have been found to be related to aerobic capacity and muscle cross-sectional area in patients with CKD predialysis and ESRD (21, 25). The current results are in agreement with previous findings on STS and CKD in demonstrating an inverse moderately significant relationship between eGFR and STS. The present data extend previous findings, showing inverse, moderately-to-highly signficant associations between RFD at time points 0-50 ms, 50-100 ms, and 0-300 ms, and the STS. Moreover, the relationship between RFD and STS was maintained when controlling for maximal force (i.e., MVF) and muscle quality (i.e., RF GSL).

Limited data are currently available regarding the impact of CKD and ESRD on RFD. Molsted et al. reported that, in dialysis patients, 16-weeks of resistance exercise training increased electromyographic (EMG) amplitude during the 200-300 ms time period following the onset of muscle contraction, with a concomitant increase of 21-38% in RFD of the knee extensors (22). These findings support resistance exercise as a possible treatment option for enhancing RFD in patients with ESRD. Gollie et al. recently highlighted the relative lack of information on the effects of resistance exercise for improving neuromuscular power, as compared to strength and muscle hypertrophy, in patients with CKD predialysis (5). Therefore, additional research is necessary to determine which resistance exercise approaches are most effective for improving RFD and physical function in such patients, and the feasibility and tolerability of such approaches.

Although peak knee extensor force (i.e., MVF) was not significantly related to functional ability in the current study, it was positively associated with absolute RFD at time points 50-100 ms, 100-200 ms, and 0-300 ms, suggesting that peak isometric knee extensor force has important implications for RFD in adults with CKD predialysis. It has been suggested that voluntary RFD becomes increasingly more dependent on maximal force generating capacity and less dependent on muscle twitch characteristics with increasing time after the onset of contraction (2, 3, 20, 27). At time intervals later than 90 ms from the onset of contraction, maximal strength accounted for 52-81% of the variance in RFD. In contrast, in very early time intervals (< 40 ms), RFD was only moderately associated with twitch contractile properties, and was less related to maximal force capacity (2).

Research on neuromuscular properties in patients with kidney disease has primarily focused on muscle size in the ESRD population on dialysis, due to their increased risk for accelerated muscle loss (34). Johansen et al. demonstrated that decreased gait speed was associated with loss in muscle contractile area and maximal isometric knee extensor strength in ambulatory hemodialysis participants (13). Conversely, John et al. reported that change in muscle cross-sectional area over 2-years was poorly correlated with change in STS (14). In non-dialysis patients, Wilkinson et al. found that muscle quality was inversely related to functional outcomes but that RF cross-sectional area was a better predictor of functional performance when compared to muscle quality (35). Our results posit RFD as an additional variable to aid in understanding the effects of of CKD predialysis on physical function and neuromuscular capacity. The lack of consistency across studies highlights the complexity of neuromuscular complications associated with CKD and the need for further research designed to clarify the relationships between neuromuscular properties and physical function in this patient population.

### Limitations

The small sample size and lack of women participants precludes generalization of the current findings beyond our study sample. The absence of control study participants, i.e. individuals without moderate to severe declines in kidney function, limits the ability to determine if, or the extent to which, RFD was compromised because of CKD in our patient population. The instructions to participants in the present study to contract as “hard” as possible may have resulted in an underestimation of RFD capabilities. Lastly, the presence of multiple comorbidities is common in patients with CKD predialysis; consequently, it is unclear how the type and number of comorbidities in our study population may have influenced the findings of the current study.

## Conclusions

In community-dwelling, middle-aged and older men with CKD stages 3 and 4, RFD is significantly associated with MVF, muscle quality and physical function. Further studies are warranted to determine the responsiveness of RFD to exercise interventions targeting physical function in patients with CKD predialysis and ESRD.

## Data Availability

De-identified, anonymized dataset will be created and shared upon the completion of the study protocol in accordance with VA policy.

## Acknowledgments

This study was conducted as part of a VA funded Center of Innovation award (VACI# AM-251) between the San Francisco VAMC and the Washington D.C. VAMC, the aim of which was to investigate clinically viable approaches for the assessment of sarcopenia in Veterans with CKD stages 3 and4 predialysis (ClinicalTrials.gov NCT03160326).

